# Genome-wide association study for circulating metabolic traits in 619,372 individuals

**DOI:** 10.1101/2024.10.15.24315557

**Authors:** Ralf Tambets, Jaanika Kronberg, Adriaan van der Graaf, Mihkel Jesse, Erik Abner, Urmo Võsa, Ida Rahu, Nele Taba, Anastassia Kolde, Dzvenymyra Yarish, Estonian Biobank Research Team, Krista Fischer, Zoltán Kutalik, Tõnu Esko, Kaur Alasoo, Priit Palta

## Abstract

Interpreting genetic associations with complex traits can be greatly improved by detailed understanding of the molecular consequences of these variants. However, although genome-wide association studies (GWAS) for common complex diseases routinely profile 1M+ individuals, studies of molecular phenotypes have lagged behind. We performed a GWAS meta-analysis for 249 circulating metabolic traits in the Estonian Biobank and the UK Biobank in up to 619,372 individuals, identifying 88,604 significant locus-metabolite associations and 8,774 independent lead variants, including 987 lead variants with a minor allele frequency less than 1%. We demonstrate how common and low-frequency associations converge on shared genes and pathways, bridging the gap between rare-variant burden testing and common-variant GWAS. We used Mendelian randomisation (MR) to explore putative causal links between metabolic traits, coronary artery disease and type 2 diabetes (T2D). Surprisingly, up to 85% of the tested metabolite-disease pairs had statistically significant genome-wide MR estimates, likely reflecting complex indirect effects driven by horisontal pleiotropy. To avoid these pleiotropic effects, we used *cis*-MR to test the phenotypic impact of inhibiting specific drug targets. We found that although plasma levels of branched-chain amino acids (BCAAs) have been associated with T2D in both observational and genome-wide MR studies, inhibiting the BCAA catabolism pathway to lower BCAA levels is unlikely to reduce T2D risk. Our publicly available results provide a valuable novel resource for GWAS interpretation and drug target prioritisation.

## Introduction

Systematic mapping and interpretation of the heritable determinants underlying complex traits and disease predisposition can be greatly improved by detailed understanding of molecular consequences of genetic variation. Studying metabolic traits is crucial as they serve as key indicators of various biological processes and disease states. Metabolite studies can reveal the complex interactions between genes and metabolic pathways, providing a more comprehensive understanding of human molecular biology and the potential for novel therapeutic hypotheses. This understanding can lead to the identification of new biomarkers for disease diagnosis, prognosis, and treatment, as well as the development of improved personalised medical interventions.

Although genome-wide association studies (GWAS) for several traits and diseases now exceed the sample size of 1 million individuals (*1–6*), studies of molecular traits such as gene expression (*7*), plasma proteins (*8*) or circulating metabolites have lagged behind. Notable exceptions are five blood lipid traits with the latest meta-analysis including data from 1.65 million individuals (*9*). Therefore, as expected, recent large-scale GWAS studies of metabolic traits continue to uncover novel associations and biological insights (*10–15*). However, for more than half of the metabolites captured by NMR, the proportion of heritability explained by genome-wide significant variants remains below 50% (*13*), indicating that much larger sample sizes are needed to discover the remaining genetic effects. Furthermore, existing GWAS studies using the Nightingale Health NMR platform have been limited to common variants (MAF > 1%) due to limited sample sizes as well as lower imputation accuracy of low frequency variants (*10–13*). Therefore, limited attention has been paid to low-frequency and rare variation, which, while explaining less heritability overall, can still provide novel biological insights (*14, 16*).

At the same time, larger sample sizes and increased statistical power of metabolic GWAS studies also brings new challenges for effectively integrating these results with GWAS findings for other complex traits and diseases. Owing to their relative simplicity, genome-wide Mendelian randomisation (MR) studies have recently gained popularity (*17–19*). Similarly to the established causal role of circulating LDL cholesterol on coronary artery disease (*20*), these studies seek to determine metabolic traits (exposures) that have a causal effect on any number of outcomes (often complex diseases). However, inferences from MR studies are only valid if certain assumptions are met (*21, 22*). A key assumption of MR is that the genetic variants are associated with the outcome only via the exposure of interest (*23*). In practice, this assumption can be challenging to satisfy, because genetic variants can have pleiotropic effects on multiple metabolic traits (*10–12, 24*). As a result, genome-wide MR studies often identify hundreds or even thousands of associations that can be challenging to interpret causally (*10, 14, 25*). A promising alternative to genome-wide MR is *cis*-MR that focuses the analysis to a specific *cis* region around the target gene of interest (*11, 26*). While *cis*-MR is less susceptible to horizontal pleiotropy (*15, 21*), it is limited by the number of independent association signals that can be identified at any gene region, thus requiring very well powered GWAS studies.

Here, we present a genome-wide association study for 249 circulating metabolic traits quantified by nuclear magnetic resonance spectroscopy (NMR) across the complete set of UK Biobank participants (n = 434,020) from diverse genetic ancestry groups, and European-ancestry individuals from the Estonian Biobank (n = 185,352) (**Figure 1**). We also performed two separate meta-analyses across the predominantly European ancestry samples (n = 599,249) and across all samples (n = 619,372), resulting in 3-5× larger sample size compared to previous studies (*10–13*) and 40% larger sample size compared to a parallel study (*14*). To illustrate the value of our resource for interpreting complex disease associations, we performed systematic genetic colocalisation between GWAS signals for coronary artery disease (CAD) and type 2 diabetes (T2D), all metabolic trait associations from this study, and gene expression and splicing QTLs from the eQTL Catalogue (*27, 28*). Finally, we demonstrate how leveraging our data in the *cis-*MR framework can be used to test specific therapeutic hypotheses. We find that although plasma levels of branched-chain amino acids (BCAAs) have been associated with T2D in both observational and genome-wide Mendelian randomisation studies (*29–32*), lowering plasma levels of BCAAs by targeting the BCAA catabolism pathway is unlikely to reduce T2D risk. Our publicly available results will provide a valuable resource for GWAS interpretation and target prioritisation studies.

**Figure 1.**
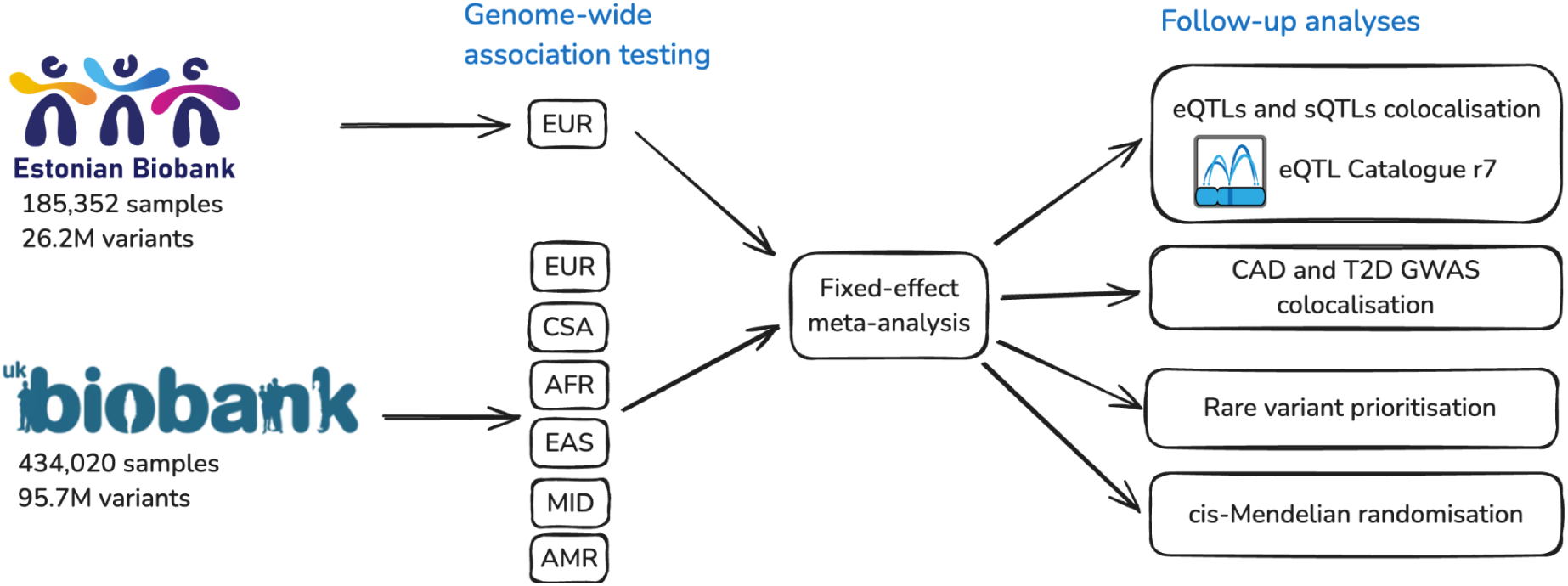
Outline of the study. First, a separate GWAS was performed for each metabolite in the Estonian Biobank and six genetic ancestry groups of the UK Biobank: EUR (European), AFR (African), AMR (Admixed American), MID (Middle Eastern), EAS (East Asian), CSA (Central/South Asian) as defined by Pan-UKBB (*33*). These ancestry-specific GWAS results were then combined in a fixed-effect meta-analysis. Finally, several follow-up analyses were performed to demonstrate the value of the resource. eQTL - gene expression quantitative trait locus; sQTL - splicing quantitative trait locus; CAD - coronary artery disease; T2D - type 2 diabetes.

## Results

### Association testing and meta-analysis

We performed GWAS for 249 metabolic traits (**Table S1**) in the Estonian Biobank (EstBB) and six genetic ancestry groups from the UK Biobank (UKBB) (**Figure 1**). The UKBB genetic ancestry groups were defined previously by the Pan-UKBB project (*33*). Relying on the population-specific genotype imputation panel for the EstBB (*34*) and the Genomics England (*35*) and the TopMed (*36*) imputation panels for the UKBB allowed us to test 10-96 million variants across genetic ancestry groups (up to 9× more than previous studies using the same NMR platform (*10, 13*)). The number of genome-wide significant (p < 5×10^-8^) locus-trait pairs ranged from 37 (UKBB_AMR) to 63,147 (UKBB_EUR) and the number of independent lead variants (r^2^ < 0.8) ranged from 24 to 6,410, with most associations detected in the UKBB_EUR and EstBB subsets (**Table 1**). Using LD score regression (*37*), we observed high genetic correlation for matched metabolic traits between the EstBB (n = 185,352) and the UKBB_EUR (n = 413,897) subsets (median rg = 0.91, mean rg = 0.89), indicating that genetic effects are largely shared between the two biobanks (**Table S2**).

**Table 1.** Number of significant locus-metabolic trait pairs (p < 5×10^-8^) and unique lead variants (r^2^ > 0.8) detected in each genetic ancestry group and the two meta-analyses.

| Biobank / genetic ancestry group | Locus-trait pairs ($p < 5 \times 10^{-8}$) | Unique lead variants | Sample size |
| --- | --- | --- | --- |
| EstBB | 27,002 | 2,356 | 185,352 |
| UKBB_EUR (European) | 63,147 | 6,410 | 413,897 |
| UKBB_CSA (Central/South Asian) | 1,070 | 112 | 8,652 |
| UKBB_AFR (African) | 916 | 143 | 6,439 |
| UKBB_EAS (East Asian) | 358 | 40 | 2,604 |
| UKBB_MID (Middle Eastern) | 92 | 44 | 1,500 |
| UKBB_AMR (Admixed American) | 37 | 24 | 928 |
| meta_EUR | 87,373 | 8,641 | 599,249 |
| meta_ALL | 88,604 | 8,774 | 619,372 |

Motivated by the high genetic correlation between the two biobanks, we proceeded with the meta-analyses. In the meta-analysis of EstBB and UKBB_EUR (meta_EUR, n = 599,249), we identified 87,373 locus-trait pairs, corresponding to 8,641 independent lead variants (r^2^ < 0.8). This represented an approximately 10-fold increase compared to the previous study by Karjalainen *et al* (*10*) (n = 136,016, 8,578 locus-trait pairs) and a 66% increase compared to a parallel study performed by Zoodsma *et al* (*14*) on the overlapping set of UKBB samples (n = 450,016, 52,662 locus-trait pairs). The estimated heritability of individual metabolic traits ranged from 2.8% for Acetoacetate to 19.5% for HDL_size (median 10.2%) and we observed a clear linear relationship between heritability and the number of loci associated with each metabolic trait (**Figure S1**, **Table S3**). On average, 95% of the lead variant associations detected by Karjalainen *et al* (*10*) also replicated in our meta_EUR analysis with a highly concordant direction of effect (**Figure S2**). We also detected many novel associations for all tested metabolites. The fraction of novel associations ranged from 27% for 3-Hydroxybutyrate (bOHbutyrate) to 85% for Lactate (**Figure 2A**). Altogether, we identified 4,085 novel independent lead variants (r^2^ < 0.8) not previously reported by Karjalainen *et al* (*10*), including 248 lead variants on the X chromosome.

**Figure 2.**
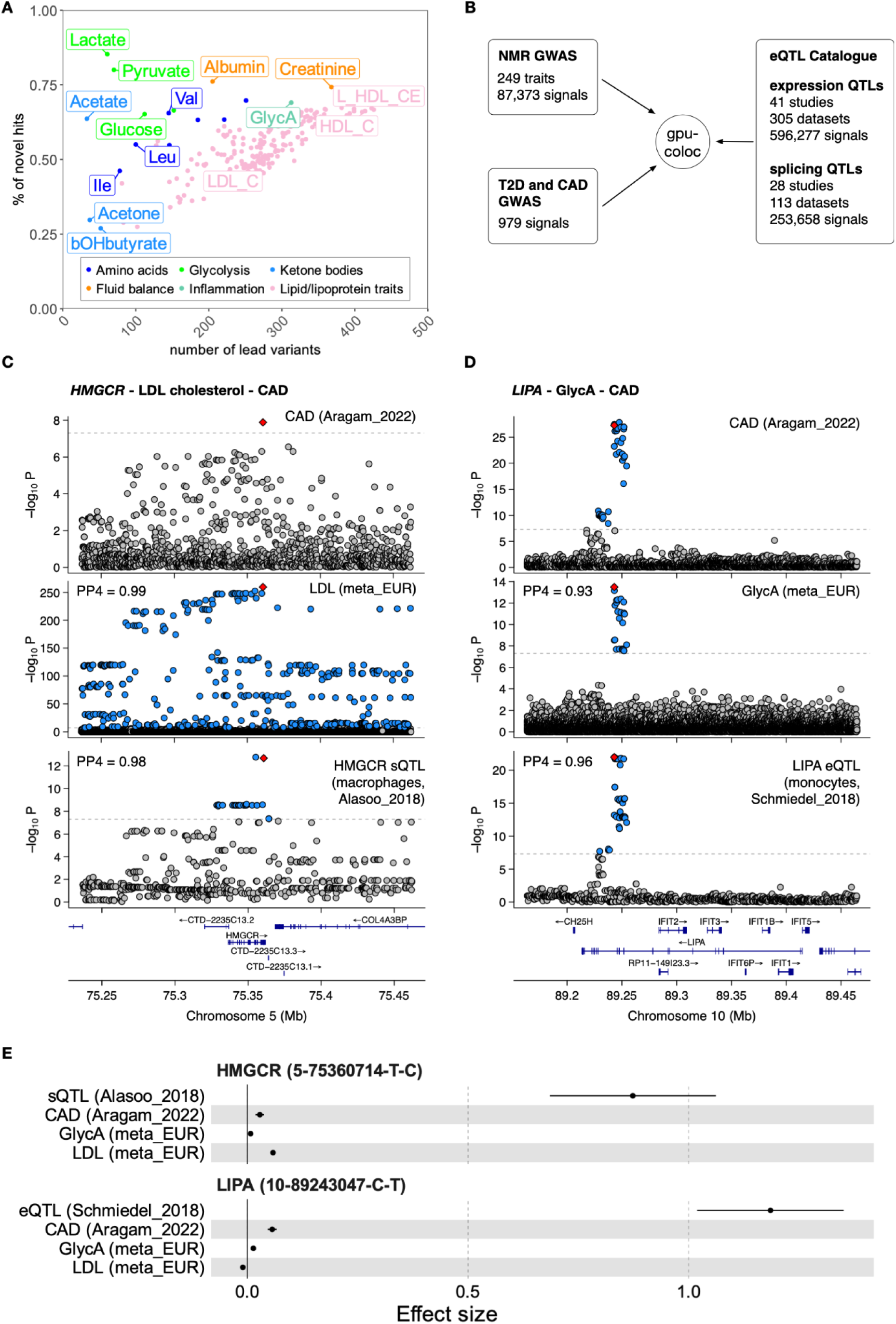
Known and novel genetic associations with metabolic traits. (**A**) Number of genome-wide significant loci (p < 5×10^-8^) detected for each metabolic trait and the proportion of those associations that were not detected by Karjalainen *et al*. (**B**) Overview of the datasets included in the colocalisation analysis. (**C**) Regional association plots for CAD, GlycA, LDL cholesterol and *HMGCR* exon 13 skipping sQTL at the *HMGCR* locus. PP4 values show pairwise colocalisation posterior probabilities between CAD and the other three traits. (**D**) Regional association plots for CAD, GlycA, LDL cholesterol and *LIPA* gene expression at the *LIPA* locus. (**E**) Effect sizes and 95% confidence intervals of the *HMGCR* and *LIPA* lead variants on CAD risk, LDL_C, GlycA and transcriptomic traits.

In addition to the EUR genetic ancestry group, we also performed GWAS in five smaller genetic ancestry groups of the UK Biobank (AFR, AMR, CSA, EAS, MID) (**Table 1**).

Including these summary statistics into our meta-analysis (meta_ALL) increased the number of independent lead variants from 8,641 to 8,774 (**Table 1**), 41 of which were not tested in the EstBB and UKBB_EUR cohorts due to low allele frequency (allele count < 20). This highlights the need to substantially increase the sample sizes for under-represented genetic ancestry groups to enable the discovery of ancestry-specific associations.

### Systematic colocalisation across molecular layers to interpret disease associations

To demonstrate how genetic associations with metabolic traits can help interpret disease associations, we colocalised all 87,373 signals from our meta_EUR analysis with quantitative trait loci (QTL) detected for gene expression (eQTLs, n = 596,277 signals) and splicing (sQTLs, n = 253,658 signals) in the eQTL Catalogue release 7 (*27, 28*), as well as GWAS signals for coronary artery disease (CAD) and type 2 diabetes (T2D) from two recent well-powered GWAS studies (n = 979 associations) (*1, 5*) (**Figure 2B**). This analysis was made feasible by our GPU-accelerated re-implementation of the coloc algorithm (*38*), leading to up to 1000-fold speed-up compared to the original R implementation (see **Methods**). Colocalisation with eQTLs and sQTLs linked 32,115 metabolic trait signals to 3,238 unique genes. Restricting the analysis to metabolic trait signals that also colocalised with T2D and CAD revealed 270 and 126 unique genes, respectively.

As expected, we detected several known gene-metabolite-disease relationships. For example, a splicing QTL affecting the inclusion of exon 13 of 3-hydroxy-3-methylglutaryl-CoA reductase (*HMGCR*) colocalised with both CAD and LDL cholesterol as well as 127 other metabolic traits from our analysis (**Figure 2C, Table S4**). *HMGCR* is a known target for statin therapy to lower circulating LDL levels and reduce CAD risk (*39*). The strongest sQTL signal for *HMGCR* was detected in the Alasoo_2018 (*40*) macrophage dataset (**Figure S3),** where statistical fine-mapping prioritised 5-75355259-A-G as the most likely causal variant (posterior inclusion probability (PIP) = 0.64), a finding that has also been validated experimentally (*41*). Consistent with a recent report, we did not detect colocalisation between the *HMGCR* sQTL signal and T2D, suggesting that T2D association at this locus involves additional genetic mechanisms (*42*). Similarly, a known fine-mapped liver-specific eQTL for *SORT1* (1-109274968-G-T, PIP = 0.99) (*43*) colocalised with LDL cholesterol and CAD risk (**Figure S4**). Also, a known pancreatic islet-specific enhancer variant 3-123346931-A-G (*44*) was specifically detected as an eQTL in the PISA pancreatic islet dataset (*45*) for *ADCY5* and colocalised with both glucose and T2D (**Figure S4**).

While focusing on novel associations, we detected a colocalisation between an eQTL signal for *LIPA* (10-89243047-C-T), CAD and 11 metabolic traits (**Table S5**). The strongest metabolic association was detected for an inflammatory biomarker GlycA (**Figure 2D**).

Consistent with a recent report (*46*), the *LIPA* eQTL signal was strongest in CD14+ monocytes and replicated across multiple independent monocyte datasets (**Table S5**). Furthermore, the GWAS effect size for GlycA was 4.3x times smaller than for CAD (**Figure 2E**), indicating that GlycA is unlikely to be a causal mediator between *LIPA* expression and CAD risk. Instead, GlycA probably represents a biomarker of an underlying inflammatory process (*47*) that drives CAD risk. This is consistent with existing literature that *LIPA* is expressed in atherosclerotic plaque macrophages, where it might contribute to atherosclerotic inflammation (*46, 48*). Thus, our results demonstrate how systematic colocalisation leveraging well-powered studies across molecular layers (RNA splicing, gene expression, metabolic traits) can aid disease GWAS interpretation and causal hypothesis generation.

### Associations with low-frequency and rare variants

While previous GWAS studies of metabolic traits profiled with NMR have primarily focused on common variation (MAF > 1%) (*10, 13, 14*), we tested all variants with minor allele count greater than 20. Thus, in our meta_EUR meta-analysis, 11.4% of the independent lead variants (n = 987 variants) had MAF < 1% (**Figure S5**). As expected, these low-frequency and rare variants also had larger effect sizes than those at higher allele frequencies (**Figure S5**). This estimate is a lower bound as many loci with common lead variants are likely to harbour secondary causal variants with lower allele frequencies.

Low-frequency variants have less extensive LD with neighbouring variants. We hypothesised that this should simplify causal gene prioritisation, because the lead variants should be more enriched for causal variants. To test this, we used computational variant effect prediction tools (*49–51*) to prioritise 63 low-frequency (MAF < 1%) missense or splice-altering variants (see **Methods**). Reassuringly, 31/63 genes with low-frequency missense or splice variants in our analysis had already been annotated in the GWAS Catalog to be associated with the same or a related trait (**Table S6**). These included low-frequency variants in *PCSK9* and *APOC3* genes associated with lipid/lipoprotein traits, a missense variant in phenylalanine hydroxylase (*PAH*) associated with phenylalanine, and a missense variant in histidine ammonia lyase (*HAL*) associated with histidine, among many others (**Table S6**). However, 32/63 of our prioritised variants were in novel genes. For example, we detected an association between the X-24503382-A-G (rs138321172) missense variant in pyruvate dehydrogenase kinase 3 (*PDK3*) and pyruvate. The *PDK3* association had likely been missed by previous studies, because the X chromosome is often excluded from GWAS studies.

As another example of novel genes not previously reported in the GWAS Catalog, we observed convergence of common and low-frequency variants on the BCAA catabolism pathway. The first two steps of BCAA catabolism are transamination of valine, leucine and isoleucine catalyzed by branched-chain aminotransferase (encoded by *BCAT1/2* genes) followed by oxidative decarboxylation catalyzed by the branched-chain α-keto acid dehydrogenase (BCKDH) complex (**Figure 3A**) (*52*). The BCKDH complex is made up of three proteins: E1 subunit encoded by the branched chain keto acid dehydrogenase E1 subunit alpha and beta genes (*BCKDHA, BCKDHB*), E2 subunit encoded by dihydrolipoyl transacylase (*DBT*) and E3 subunit encoded by dihydrolipoamide dehydrogenase (*DLD*) (*52*) (**Figure 3B**). The activity of the *BCKDH* complex is further regulated by the branched chain keto acid dehydrogenase kinase (*BCKDK*) that inhibits its activity, and protein phosphatase 2Cm (encoded by *PPM1K)* that reactivates it (**Figure 3B**).

**Figure 3.**
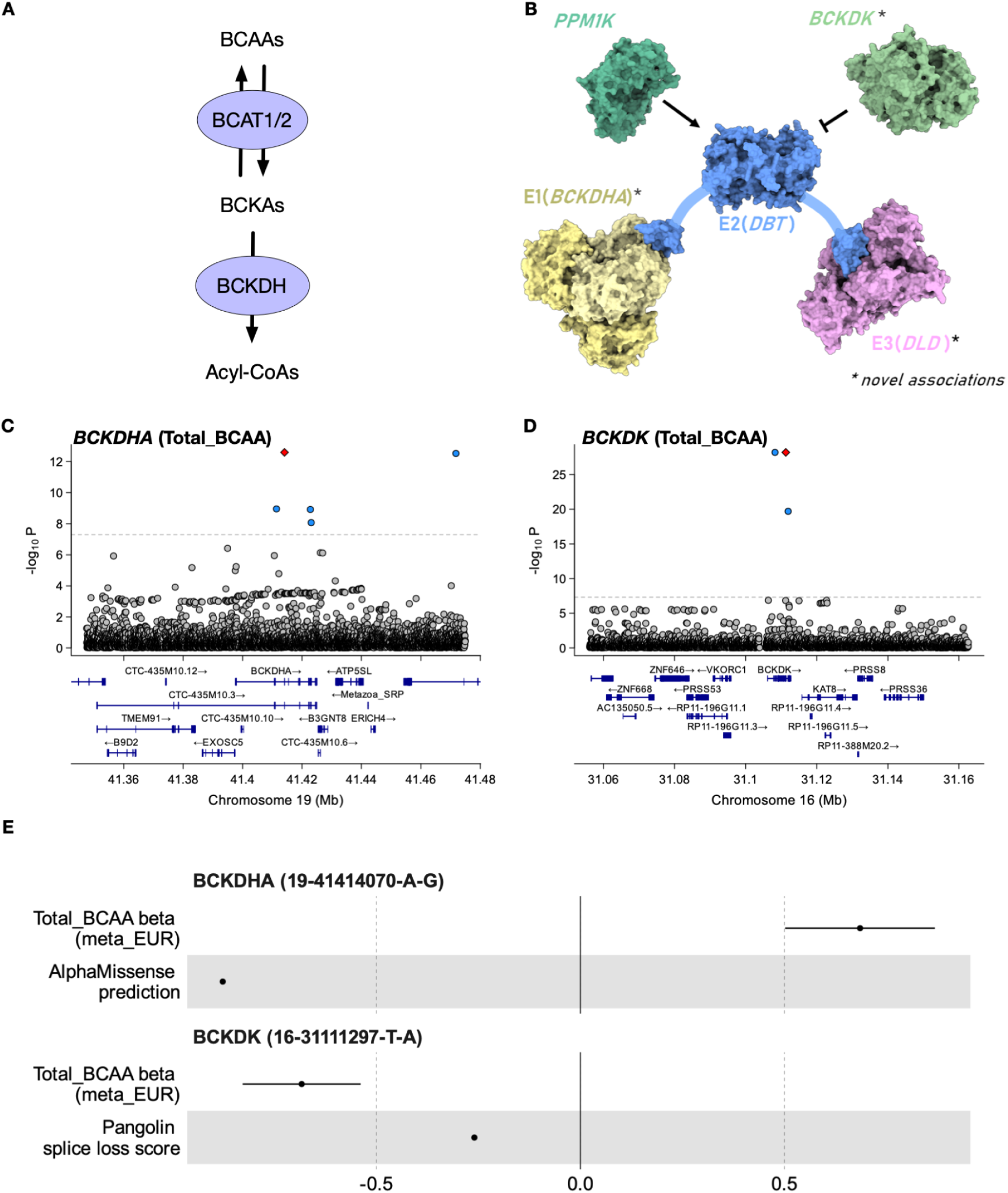
Convergence of common and low-frequency associations at the branched-chain amino acid catabolism pathway. (**A**) Branched-chain amino acids (BCAAs) are converted to branched chain keto acids (BCKAs) by branched-chain amino acid aminotransferase (encoded by *BCAT1* and *BCAT2*). This process is reversible. BCKAs can be further catabolised by the branched-chain α-keto acid dehydrogenase (BCKDH) complex to acyl-CoAs. (**B**) BCKDH complex is made up of subunits E1 (encoded by *BCKDHA* and *BCKDHB)*, E2 encoded by *DBT* and E3 encoded by *DLD*. The activity of the BCKDH complex is controlled by a kinase (*BCKDK*) that inhibits its function, and phosphatase (*PPM1K*) that reactivates it (*52*). (***C***) GWAS association signal for Total_BCAA in the *BCKDHA* gene region. The *BCKDHA* missense variant 19-41414070-A-G is highlighted in red. (**D**) GWAS association signal for Total_BCAA in the *BCKDK* gene region. The predicted *BCKDK* splice loss variant 16-31111297-T-A is highlighted in red. (**E**) The effect of the *BCKDHA* missense variant and *BCKDK* splice loss variant on Total_BCAA (beta + 95% confidence interval) together with variant effect predictions on protein function from AlphaMissense (*53*) and Pangolin (*51*) models (arbitrary units).

Common-variant associations for three of the six genes (*BCAT2*, *DBT*, and *PPM1K*) have been reported in previous GWAS studies for BCAAs. We additionally detected a low frequency (MAF = 0.012%, p = 2.6×10^--13^) missense variant 19-41414070-A-G (rs771686663) in *BCKDHA* (**Figure 3C)** and a low frequency (MAF = 0.047%, p = 6.5×10^-29^) splice region variant 16-31111297-T-A (rs118042732) in *BCKDK* (**Figure 3D)**. In both cases, the predicted variant effects were directionally consistent with the sign of the GWAS associations (**Figure 3E**). The *BCKDHA* missense variant was predicted by CADD (*54*) and AlphaMissense (*53*) to be deleterious and was associated with increased BCAA levels. In contrast, the rs118042732 splice region variant was predicted by Pangolin deep learning splice site prediction model (*51*) to lead to splice acceptor loss and was associated with decreased BCAA levels (consistent with BCKDK being a negative regulator of the BCKDH complex) (**Figure 3B**). Reassuringly, both *BCKDK* (p < 1×10^-30^) and *BCKDHA* (p < 1×10^-13^) were also found to be associated with BCAAs in a parallel effort that performed rare-variant burden testing and exome-wide association testing using overlapping UK Biobank NMR samples (*14*), confirming that our rare variant imputation is reliable.

Finally, we detected a novel common-variant (7-107837919-T-A, MAF ∼50%) association at the *DLD* locus (beta = -0.01, p = 9.8×10^-13^). Thus, we identified GWAS hits for all six key enzymes involved in the catabolism of BCAAs. This illustrates that very large sample sizes are needed to saturate the discovery of key regulators of biological processes due to either very small effects of some common variants on the target genes (*DLD*) or very low allele frequency of the genetic variants that affect those genes (*BCKDHA*, *BCKDK*). Interestingly, while the GWAS and burden testing analysis by Zoodsma *et al* (*14*) identified largely divergent set of genes in this pathway (*BCAT2*, *BCKDK* and *BCKDHA* from burden testing and *BCAT2*, *DLD*, *DBT* and *PPM1K* from GWAS), our well-powered GWAS using imputed low frequency variants was able to discover all six genes in a single analysis. This is consistent with recent reports that the differences between GWAS and burden testing results can be largely explained by differential statistical power (*55*).

### Extent of horizontal pleiotropy across metabolic traits

To understand the shared genetic control of various classes of metabolic traits, we explored genetic correlations between all 249 metabolic traits (*37*). Although the median genetic correlation across all traits was low (rg = 0.16), there were high genetic correlations between various lipoprotein traits (median rg = 0.52) as well as between other closely regulated metabolites, such as BCAAs (rg = 0.97) (**Figure S6**, **Table S7**). To better understand the molecular mechanisms behind genetic correlations, we identified clusters of lead variants that were shared (r^2^ > 0.8) between metabolic traits. Among the 249 metabolic traits, most lead variants were significantly associated (p < 5×10^-8^) with multiple metabolites (mean = 10; median = 2). Most prominently, a common missense variant (MAF = 40%) in the glucokinase regulatory protein (*GCKR*) (2-27508073-T-C, *GCKR*:p.Leu446Pro) was significantly associated (p < 5×10^-8^) with 231 (out of 249) metabolites (**Figure 4A**). However, in many other cases, pleiotropy was restricted to the same class of metabolites such as the 5-75355259-A-G variant at the *HMGCR* locus with multiple lipid traits (**Figure S7**).

**Figure 4.**
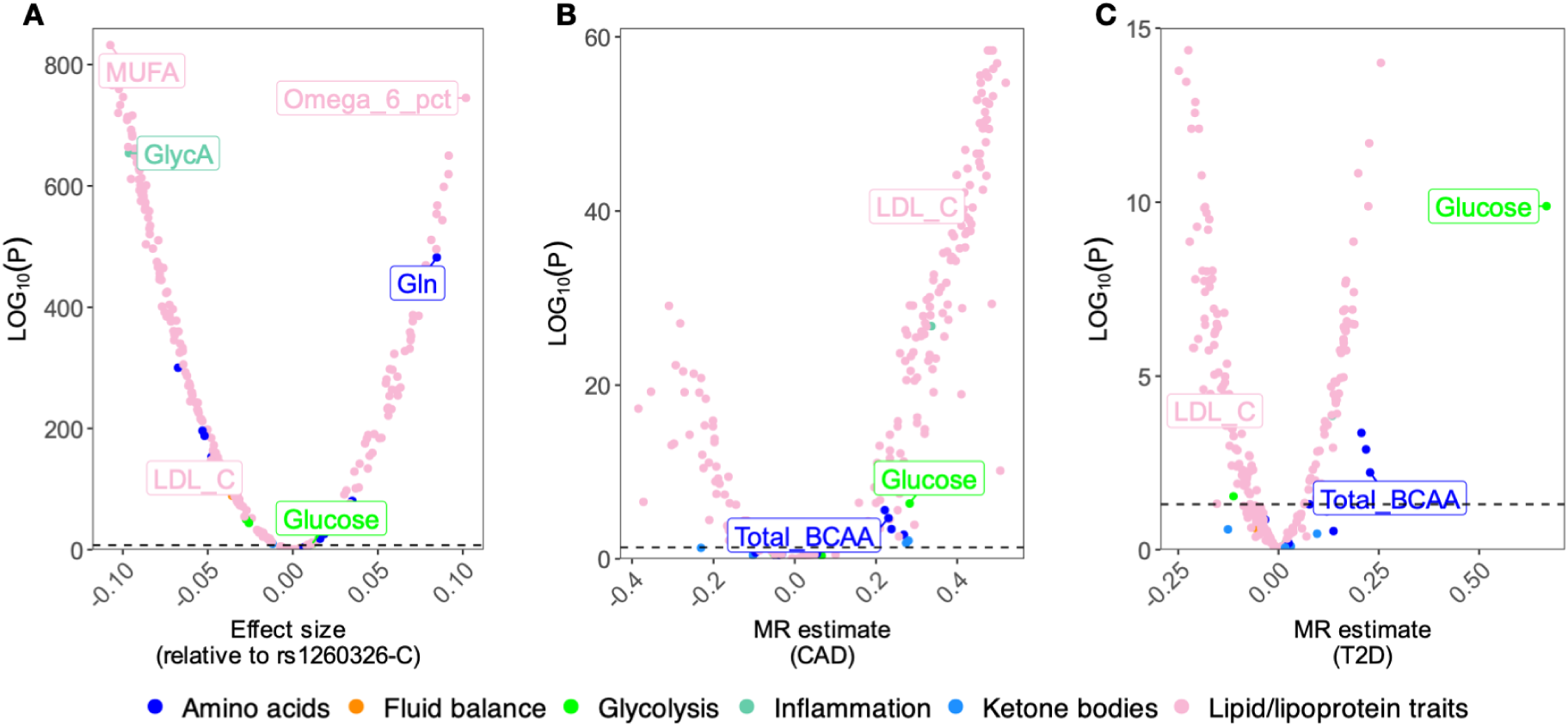
Extent of pleiotropic associations across metabolic traits. (**A**) Pleiotropic effects of the *GCKR* missense variant 2-27508073-T-C (rs1260326) on 249 metabolites. (**B**) Genome-wide MR estimates using all 249 metabolic traits as exposures and CAD as outcome. (**C**) Genome-wide MR estimates using all 249 metabolic traits as exposures and T2D as outcome.

To characterise the impact of pleiotropic genetic effects on interpreting disease associations, we performed genome-wide Mendelian randomisation (MR) using all 249 metabolic traits as exposures and either coronary artery disease (CAD) (*5*) or type 2 diabetes (T2D) (*1*) as outcomes (see Methods). For CAD, 211 of the 249 (85%) tested metabolic traits yielded significant MR estimates (FDR < 5%, **Figure 4B**) while for T2D (**Figure 4C**) the number of significant associations was 157 (63% of tested traits) (**Table S8**). Reassuringly, we recapitulated known causal effects between genetically regulated LDL cholesterol and CAD (beta = 0.43, p-value = 6.02×10^-42^) and between glucose and T2D (beta = 0.67, p-value = 6.3×10^-12^). We also detected a known negative association between genetically lower LDL cholesterol and T2D (beta = -0.11, p-value = 2.37×10^-4^) (*56*). Finally, we detected genome-wide significant MR estimates between BCAA levels and both CAD and T2D

(**Figure 4B-C**). However, these genome-wide MR estimates, beyond the known causal effects of LDL and glucose, can be tricky to interpret due to extensive genetic correlation between the metabolic traits (**Figure S6, Table S7**), widespread horizontal pleiotropy, and large heterogeneity between the effect estimates from individual variants (**Table S8**).

### Evaluating drug targets with *cis*-Mendelian randomisation

To limit the impact of horizontal pleiotropy, we interrogated a subset of the genome-wide MR associations using a more conservative *cis*-MR (drug-target MR) approach (*21, 26*). Instead of capturing average genome-wide effects of circulating metabolic traits, *cis*-MR uses genetic variation in *cis* of a drug target gene to estimate the impact of perturbing gene function on disease risk (*26*). If these genes have a direct biological effect on metabolic traits, then we can use the variant effect on those traits as a proxy read-out for the (unmeasured) effect of these variants on gene function (*15*). First, we focussed on three genes with direct effect on regulating plasma LDL cholesterol levels: low density lipoprotein receptor (*LDLR*), *HMGCR*, and proprotein convertase subtilisin/kexin type 9 (*PCSK9*) (**Figure 5A**). In all three cases, we observed robust causal effects of perturbing these genes on CAD risk as previously reported (*11, 56*). We then estimated the causal effect of lowering LDL cholesterol via these mechanisms on T2D. At the *HMGCR* locus, we detected a negative association between genetically regulated LDL cholesterol and T2D, which is consistent with previous MR studies as well as large clinical trials demonstrating that statin use is associated with increased T2D risk (*42, 56*). Notably, while the effect of genetically regulated LDL cholesterol on CAD risk was even higher at the *LDLR* and *PCSK9* loci, the effect on T2D was strongly attenuated relative to *HMGCR* (*56*). This is consistent with clinical trials of *PCSK9* inhibitors not detecting increased risk of T2D as a side effect (*57*).

**Figure 5.**
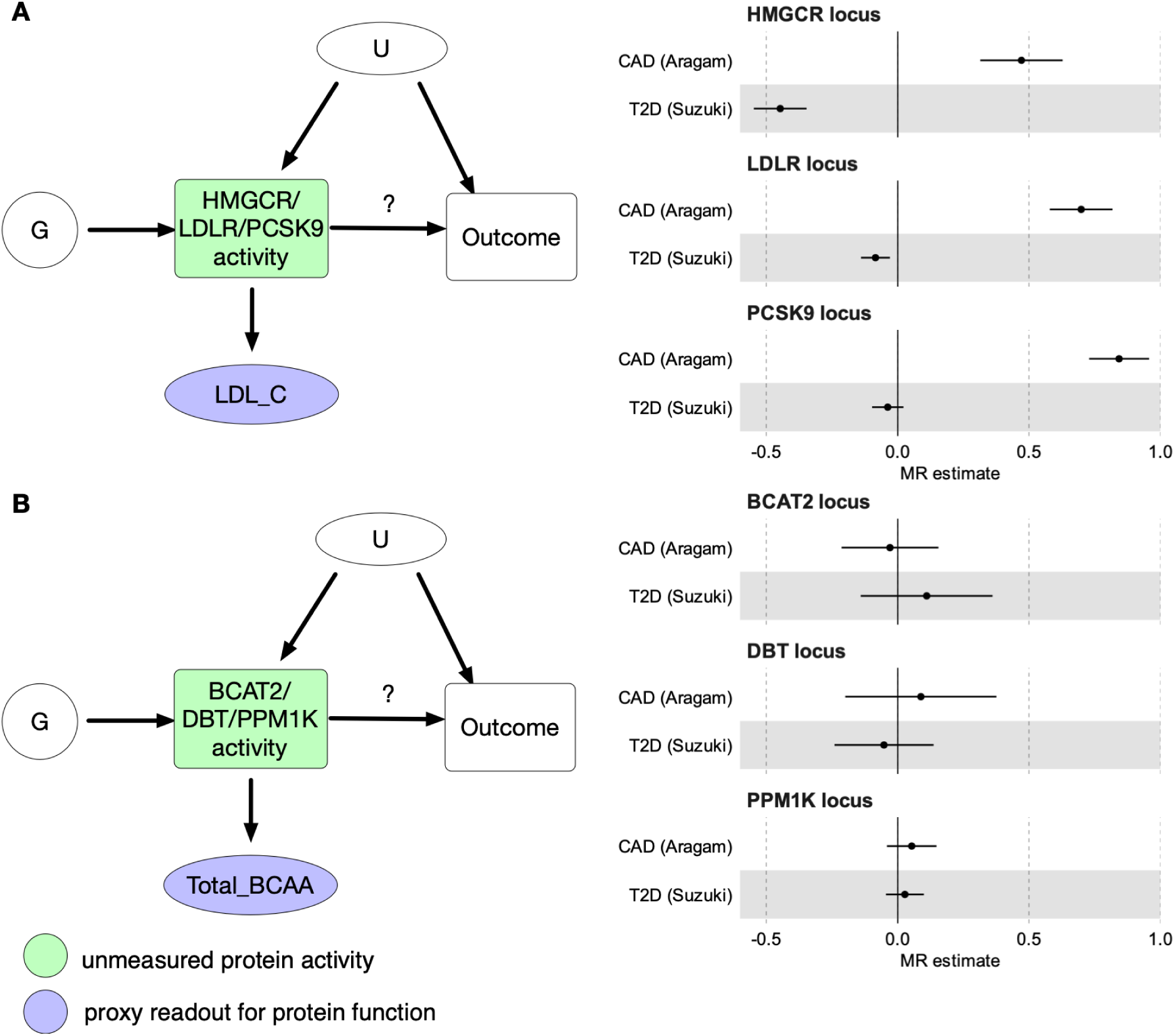
Drug target evaluation with *cis*-Mendelian randomisation. **(A)** Performing *cis*-MR using genetic variants G from the *cis* regions of *HMGCR*, *LDLR* and *PCSK9* genes to estimate the causal effect of inhibiting the corresponding gene function on T2D and CAD risk. LDL cholesterol (LDL_C) is used as a proxy readout for variant effects on *HMGCR*, *LDLR* and *PCSK9* function. (**B**) Performing *cis-*MR using genetic variants G from the *cis* regions of *BCAT2*, *DBT* and *PPM1K* genes to estimate the causal effect of inhibiting the corresponding protein function on T2D and CAD risk. Total branched-chain amino acid levels (Total_BCAA) is used as a proxy readout for variant effect on *BCAT2*, *DBT* and *PPM1K* function. U - unmeasured confounders.

Our results illustrate how a carefully conducted *cis*-MR analysis with well-powered proxy exposures can be used to evaluate drug targets and quantify their potential side effects.

Reassured by the ability of *cis*-MR to rediscover known associations with lipid-lowering drug targets, we next followed up the significant genome-wide MR estimates between BCAAs and both CAD and T2D (**Figure 4B-C**). Although the association between genetically regulated BCAAs and T2D has been reported before (*29*), genome-wide MR necessarily averages effects across multiple distinct mechanisms, only some of which might influence T2D risk.

Furthermore, recent studies have suggested that the genome-wide MR signal between BCAAs and T2D first observed by (*29*) might be confounded by horizontal pleiotropy (*14, 58*). As discussed above, a prominent mechanism regulating plasma BCAA levels is BCAA catabolism controlled by BCAT2 and the BCKDH complex (**Figure 3A**). To clarify the contradictory results obtained from genome-wide MR analysis and motivated by the recent discovery of a clinical candidate of the BCKDK kinase inhibitor (*59*), we sought to assess if lowering plasma BCAA levels via specific inhibition of the BCKDK kinase could reduce T2D and CAD risk.

The most direct way to assess this would be to perform *cis*-MR with genetic variants in the *cis* region of *BCKDK* as genetic instruments, plasma BCAA levels as a proxy exposure and T2D as outcome. However, the *BCKDK* region lacks strong common variant associations and the low-frequency splice donor variant that we detected (**Figure 3D**) is too rare to have sufficient power for *cis*-MR. Instead, we focussed on *cis* variation near *DBT* and *PPM1K*, two other members of the BCKDH complex (**Figure 3B**) that have robust common variant associations. We also included *cis* variation near *BCAT2*, an enzyme directly upstream of BCKDH complex in the BCAA catabolism pathway (**Figure 3A**). In all three gene regions, the results were broadly consistent with a null effect of BCKDK inhibition on T2D risk (**Figure 5B**). None of these loci had genome-wide significant hits for T2D (**Figures S8-S9**). Although some MR method and outcome GWAS combinations (see **Methods**) did yield non-null causal effect estimates, these were not consistent across the three *cis* regions (**Tables S9-S11**). Thus, current genetic evidence does not support the hypothesis that *BCKDK* inhibition would have a large beneficial effect on reducing T2D risk.

One limitation of *cis*-MR is that due to the focus on a narrow *cis*-region for the selection of genetic instruments, it can be sensitive to linkage disequilibrium (LD) between independent causal variants (*60, 61*). For example, although the association signals between LDL cholesterol and T2D at the *HMGCR* locus do not colocalise with each other (*42*), the two lead variants are in moderate LD (r^2^ = 0.53), which could likely bias our *cis*-MR estimates for T2D (**Figure 2A**) at this locus. This issue of LD is likely to become more pronounced as the sample size and power of GWAS studies increase. For example, even though the lead variant at the *BCAT2* locus (19-48800958-C-T) has a genome-wide significant (p < 5×10^-8^) association with 30 lipid traits, this seems to be entirely driven by low LD (r^2^ = 0.08) with a neighbouring *FUT2* locus lead variant (19-48703417-G-A) (**Figure S8**).

## Discussion

We have created a comprehensive resource of both common and low-frequency genetic variants associated with 249 metabolic traits in up to 619,372 individuals across multiple ancestry groups. We have demonstrated the utility of the resource for GWAS interpretation via systematic phenome-wide colocalisation, low-frequency variant prioritisation and *cis*-MR analysis. To ensure that our results can be used as widely as possible, we have publicly released all summary statistics via the GWAS Catalog (*62*) and we have also made the results easy to query via an online browser (https://nmrmeta.gi.ut.ee/). Thus, our study provides the most comprehensive catalogue of genetic associations with these metabolites yet. All our analyses were performed on the latest human reference genome (GRCh38), greatly simplifying integration with other large-scale human genetics resources such as FinnGen (*63*), Million Veterans Program (*64*) and the eQTL Catalogue (*28*) without the risk of losing genetic variants through the lift-over process (*65*).

Although plasma BCAA levels have been robustly associated with T2D diabetes in observational studies (*30–32*), this effect seems to be primarily driven by reverse causality whereby variants associated with insulin resistance (*66, 67*) and T2D (*58*) increase plasma BCAA levels. Genome-wide MR studies using BCAA levels as exposure and T2D as outcome have yielded contradictory results, with our genome-wide MR and other early studies suggesting that BCAA levels increase T2D risk (*29*), but more recent studies finding that this association is significantly attenuated when pleiotropic genetic variants are excluded (*14, 58*). However, even when care is taken to exclude pleiotropic variants, genome-wide MR still averages over multiple distinct mechanisms that could alter plasma BCAA levels, only some of which could potentially have a causal effect on T2D. Here, we performed focussed *cis*-MR analysis with genetic variants from *PPM1K*, *DBT* and *BCAT2* gene regions as instruments to directly assess the potential therapeutic effect of inhibiting BCKDK kinase on reducing T2D risk. Our results were consistent with a null effect, suggesting that the recently published clinical candidate of the BCKDK kinase inhibitor (*59*) is unlikely to reduce T2D risk.

Our work is highly complementary to a parallel effort by Zoodsma *et al* (*14*) that used an overlapping set of ∼450,000 UK Biobank samples (*14*). First, including the Estonian Biobank into the meta-analysis increased the sample size by ∼40% compared to UK Biobank alone and increased the number of genome-wide significant discoveries by approximately 64%. Secondly, instead of relying on old genotype imputation from the UK Biobank for GWAS analysis (*68*), we used the latest available set of imputed genotypes from both biobanks (*34–36*). This allowed us to test ∼10x more variants in the UK Biobank and 2.5x more variants in the Estonian Biobank compared to Zoodsma *et al.* As a result, 11.4% of the independent lead variants detected in our analysis had a minor allele frequency (MAF) of less than 1% (**Figure S5**). Most notably, our analysis detected genome-wide significant hits for six members of the BCAA catabolism pathway, two of which (*BCKDHA* and *BCKDK*) were missed by the GWAS performed by Zoodsma *et al.,* due to lack of common variant associations at these loci. Reassuringly, both genes were detected by rare variant burden testing and exome-wide association analysis (*14*), confirming that our findings are reliable even at low allele frequencies. Thus, our results illustrate how well-powered GWAS studies that test low-frequency and rare variants can help to reconcile the apparent differences in gene discovery from common-variant GWAS and rare variant burden testing studies (*55, 69*).

Our study also has several limitations. First, 97% of the samples included in the analysis were of predominantly European genetic ancestries. This skew limited our ability to detect genome-wide significant signals in other genetic ancestry groups and may influence the generalizability of our findings across genetic ancestry groups. As a result, the number of genome-wide significant signals increased by only 1.4% (**Table 1**) when samples from other UKBB genetic ancestry groups (AFR, AMR, CSA, EAS, MID) were included in the meta-analysis. Secondly, due to significant methodological challenges (*70*), we did not perform statistical fine mapping of the identified loci. As a result, we are likely missing many secondary signals at the genome-wide significant loci. Lastly, we applied a global genome-wide significance threshold (p < 5×10^-8^) tailored for common variants. To control for false-positive associations, this threshold may need adjustment to account for the large number of metabolic traits tested and the low allele frequency threshold (allele count > 20) utilised in our study.

While biobank-scale datasets provide unprecedented power for genetic discovery, they also introduce complexities in interpreting genetic associations due to pervasive pleiotropy. Our results reinforce previous reports of extensive pleiotropy across metabolic trait GWASs (*10–12*). Some of this pleiotropy is readily interpretable, such as co-regulation between various lipid traits (*11*) or opposing effects between substrates and products of enzymatic reactions (*12*). However, given our large sample size, we also detected more cryptic pleiotropic effects such as the *GCKR* missense variant that was associated with 231/249 tested metabolic traits (**Figure 4A**). Such extensive overlaps exemplify that as cohorts grow larger, the detection of pleiotropic signals becomes more pronounced, making it harder to disentangle direct and indirect genetic effects. As a result, we caution against interpreting genome-wide MR results as evidence of direct causal effect of tested metabolic traits on the outcomes of interest (*23, 24*). *Cis*-MR analyses are potentially less susceptible to these pleiotropic effects, but still require careful consideration of LD and detailed understanding of metabolic pathways to ensure that MR assumptions are met (*15, 21, 26*).

## Methods

### Cohorts

#### Estonian Biobank

The Estonian Biobank (EstBB) is a population-based biobank at the Institute of Genomics, University of Tartu (*71*). The current EstBB data freeze consists of 212,955 adult (age≥18y) participants, reflecting the age, sex and geographical distribution of the adult Estonian population, for whom biological samples as well a variety of health-related and demographic information have been collected. All biobank participants have signed a broad informed consent form and their blood sample collection was undertaken across the country between 2002 and 2021 (*71, 72*). The activities of EstBB are regulated by the Human Genes Research Act, which was adopted in 2000 specifically for the operations of EstBB. Nightingale Health NMR platform was used to generate plasma metabolite profiles for all individual samples in the biobank. The assay covers 249 metabolic traits ranging from low molecular weight compounds to lipids and lipoproteins. Individual level data analysis in EstBB was carried out under ethical approval 1.1-12/624 from the Estonian Committee on Bioethics and Human Research (Estonian Ministry of Social Affairs), using data according to release application 6-7/GI/8988 from the EstBB.

#### UK Biobank

The UK Biobank is a longitudinal biomedical study of approximately half a million participants between 38-71 years old from the United Kingdom (*68*). Participant recruitment was conducted on a volunteer basis and took place between 2006 and 2010. Initial data were collected in 22 different assessment centers throughout Scotland, England, and Wales. Data collection includes elaborate genotype, environmental and lifestyle data.

Blood samples were drawn at baseline for all participants, with an average of four hours since the last meal, i.e. generally non-fasting. NMR metabolic traits (Nightingale Health, quantification library 2020) were measured from EDTA plasma samples (aliquot 3) during 2019–2024 from the entire cohort. Details on the NMR metabolomic measurements in UK Biobank have been described previously for the first tranche of ∼120,000 samples (*73*). The UK Biobank study was approved by the North West Multi-Centre Research Ethics Committee. This research was conducted using the UK Biobank Resource under application numbers 91233 and 30418.

### Genotype imputation

#### Estonian Biobank

All EstBB participants have been genotyped at the Core Genotyping Lab of the Institute of Genomics, University of Tartu, using Illumina Global Screening Array v1.0, v2.0 and v3.0. Samples were genotyped and PLINK format files were created using Illumina GenomeStudio v2.0.4. Individuals were excluded from the analysis if their call-rate was < 95%, if they were outliers of the absolute value of heterozygosity (> 3SD from the mean) or if sex defined based on heterozygosity of X chromosome did not match sex in phenotype data (*34*). Before imputation, variants were filtered by call-rate < 95%, HWE p-value < 1e-4 (autosomal variants only), and minor allele frequency < 1%. Genotyped variant positions were lifted over from GRCh37 to GRCh38 with Picard. Phasing was performed using the Beagle v5.4 software (*74*). Imputation was performed with Beagle v5.4 (beagle.22Jul22.46e.jar) using default settings. Dataset was split into batches of 5,000 variants. A population specific reference panel consisting of 2,695 WGS samples (*34*) was utilised for imputation and standard Beagle hg38 recombination maps were used. Based on principal component analysis, samples who were not of European ancestry were removed. Duplicate and monozygous twin detection was performed with KING 2.2.7 (*75*), and one sample was removed out of the pair of duplicates.

#### UK Biobank autosomes

Genotype imputation for the UK Biobank (UKBB) autosomal data was conducted using a high-coverage whole genome sequencing reference panel (342 million autosomal variants) from 78,195 individuals from the Genomics England (GEL) project. Reference panel construction and UK Biobank imputations have been described previously (UKBB data field 21008) (*35*).

Briefly, the UKBB SNP array data consisted of 784,256 autosomal variants. Initially, 113,515 sites identified by previous centralised UK Biobank analysis as failing quality control were removed, along with an additional 39,165 sites failing a Hardy–Weinberg equilibrium test on 409,703 GBR samples, with a p-value threshold of 1^-10^. The resulting SNP array data were mapped from the GRCh37 to GRCh38 genome build using the GATK Picard LiftOver tool.

Alleles with mismatching strands but matching alleles were flipped. A further 495 sites were removed due to incompatibility between the two reference genomes, resulting in a final SNP array incorporating 631,081 autosomal variants used for phasing and imputation.

Haplotype estimation of the SNP array data, a prerequisite for imputation, was carried out one chromosome at a time using SHAPEIT4 v4.2.2 (*76*) without a reference panel, utilising the full set of UK Biobank samples. SHAPEIT4 was run with its default 15 Markov chain Monte Carlo iterations and 30 threads. Autosomal imputation using the GEL reference panel was conducted with IMPUTE5 (*77*) (v.1.1.4). The SNP array data were divided into 408 consecutive and overlapping chunks of approximately 5 megabases (Mb) each, with a 2.5 Mb buffer across the genome using the Chunker program in IMPUTE5. Each chunk was further divided into 24 sample batches, each containing 20,349 samples. IMPUTE5 was run on each of the 9,792 subsets using a single thread and default settings. The resulting imputed genotype dosages are stored in BGEN format, and phasing information is stored in VCF format.

#### UK Biobank X chromosome

As the UKBB genotypes imputed by Genomics England did not include the X chromosome, we used the TOPMed r2 imputation for the X chromosome (UKBB data field 21007). Imputation was performed using the TopMed Imputation Server (*78*). The data were divided into 10 Mb chunks, and each chunk underwent several checks to ensure validity. These checks included verifying the inclusion of variants in the reference panel, ensuring a sufficient overlap with the reference panel, and maintaining an adequate sample call rate. Chunks that did not meet these criteria were excluded from further analysis. Overall, quality control methods employed by the TopMed Imputation Server were slightly more conservative than those employed by GEL and thus the sample size for each sub-population decreased by roughly 0.5% (final sample sizes: AFR - 6,411; AMR - 925; CSA - 8,627; EAS - 2,595; EUR - 412,523; MID - 1,491). Genotype phasing was performed with Eagle2 (*79*) and imputation was conducted with mimimac4 (*78*). After imputation, all chunks of each chromosome were merged into a single file. For chromosome X, additional checks were performed to verify ploidy and ensure the accuracy of mixed genotypes. The chromosome was split into three regions (PAR1, non-PAR, PAR2) for phasing and imputation, and these regions were later merged into a complete chromosome X file.

### NMR metabolite data quality control and normalisation

NMR data generation in the EstBB and UKBB has been previously described (*80*). During the quality control of the Nuclear Magnetic Resonance spectroscopy (NMR) metabolomics data, we detected a difference between distributions of several metabolites (notably Ala and His) driven primarily by spectrometer and batch effect. We removed this unwanted technical variation using the R package ’ukbnmr’ in both EstBB and UKBB data (*81*). We excluded individuals with more than 5 missing metabolite measurements from the cohort, confirmed that none of the 249 metabolites had a significant number of missing measurements (8000 for EstBB, 24000 for UKBB), and applied a metabolite-wise inverse normal transformation to obtain the final dataset.

### Association testing and meta-analysis

We conducted genome-wide association tests for each of the seven genetic ancestry groups separately using regenie v3.1.1 (*82*), with sex, age, age squared and the top principal components of the genotype data used as covariates (PC1-PC10 for EstBB, PC1-PC20 for UKBB). For step 1 (whole genome model), we used genotype calls for UKBB and genotyping data for EstBB and included variants with a minor allele frequency (MAF) of at least 1%, a minor allele count (MAC) of at least 20, Hardy-Weinberg equilibrium exact test p-values of 10^-15^ or less, and maximum per-variant and per-sample missing genotype rates of 0.1. For step 2 (association testing using a linear regression model), we used imputed genotypes and selected variants with a MAC of at least 20 and an imputation INFO score of at least 0.6.

We performed two different inverse-variance weighted fixed-effect meta-analyses: meta_EUR on individuals of predominantly European genetic ancestry (EstBB cohort and EUR genetic ancestry group of UKBB), and meta_ALL which encompasses all seven genetic ancestry groups from UKBB and EstBB.

### Genetic correlations

We employed LD score regression (LDSC) (*83*) to obtain pairwise genetic correlations for all 249 NMR metabolites. Correlations were calculated between biobanks for each metabolite and between all metabolites in three of the largest datasets (EstBB, UKBB_EUR and meta_EUR) using the European reference panel LD scores.

### Lead variant and locus definition

We obtained the set of dataset-metabolite-variant triplets by iterating over variants that met the genome-wide significance threshold of 5×10^-8^. The variant with the lowest p-value was designated as the lead variant within a 2 Mb locus. In each dataset, neighbouring loci were merged into one if their lead variants were in LD with an r^2^ of at least 0.05. To better evaluate the independence of lead variants, we utilised PLINK v1.90b6.26 to calculate pairwise LD between all lead variants in a single genetic ancestry group, assigning them into shared cross-metabolite clusters if r^2^ was at least 0.8. The variant with the smallest p-value was assigned as the lead variant for each cluster.

### Colocalisation

We performed colocalisation between all genome-wide significant locus-trait pairs from the meta_EUR meta-analysis, GWAS hits for T2D and CAD from two large meta-analyses (*1, 5*), and all gene expression and splicing QTLs from the eQTL Catalogue release 7 (*27, 28*). For metabolite and disease GWAS loci, we extracted summary statistics from the +/- 1Mb window around each lead variant and converted those to approximate Bayes factors using the formula defined in the process.dataset() function of the coloc R package (*84*). For the eQTL Catalogue datasets, we used the fine-mapped (*85*) log Bayes factors available from the eQTL Catalogue FTP Server (*28*). For eQTLs, we included all datasets from the eQTL Catalogue dataset <u>metadata table</u> where the quantification method was either ‘ge’ or ‘microarray’. For sQTLs, we included all datasets where the quantification method was ‘leafcutter’ (*86*). We then used the algorithm defined in the bf_bf() function of the coloc R package (*38*) to test for colocalisation between the GWAS approximate Bayes factors and eQTL/sQTL log Bayes factors. For prior probabilities, we used p1 = 1e-4, p2 = 1e-4, p12 = 1e-6. We re-implemented all computations in Python and used PyTorch (*87*) to add support for GPU acceleration. Parallelisation of colocalisation testing on GPUs combined with efficient storage of summary statistics in Parquet files allowed us to achieve ∼1000-fold speed-up compared to the original R implementation (*38*). Our gpu-coloc source code is available at https://github.com/mjesse-github/gpu-coloc.

### Prioritisation of low-frequency missense and splice-altering variants

We first identified all independent lead variants in the meta_EUR analysis that had MAF < 1%. We then narrowed the set down by only including SNPs identified as missense or splice regions variants by the Ensembl Variant Effect Predictor (VEP) (*49*) or were predicted to alter splice junction usage by SpliceAI (*50*) or Pangolin (*51*). This approach identified 63 variants that we were able to assign to putative effector genes (Table S6). We then downloaded all associations from the GWAS Catalog (*62*) (https://www.ebi.ac.uk/gwas/docs/file-downloads, accessed 8 February 2025) where the ‘Mapped gene’ column matched one of our 63 prioritised genes. Finally, we assessed if the traits reported in the GWAS Catalog were related to the metabolic traits detected in our analysis (Table S6).

### Genome-wide Mendelian randomisation

We performed genome-wide MR between all 249 metabolic traits and two diseases, CAD (*5*) and T2D (*1*), resulting in a total of 498 analyses. For each metabolic trait, we identified instrumental variables using a greedy LD pruning approach applied to its lead variants with MAF > 1%. This involved (A) assigning the lead with the lowest p-value in the initial set to the instrument set, (B) discarding that variant and all variants in LD with it (r^2^ < 0.01) from the initial set, and (C) repeating steps A and B until no variants remained in the initial set. For the MR analysis itself, we used multiplicative random-effects inverse-variance weighted MR (IVW-MR) (implemented in MendelianRandomization R package (*88*)) as recommended by recent best practices (*21*).

### *cis*-Mendelian randomisation

For the primary *cis*-MR analysis, we included only variants from the +/- 200kb region around the gene body of the target gene that had MAF > 1%. For instrument selection, we used the LD information from the UK Biobank Genomics England imputation (Pan-UKBB EUR subset, n = 413,897) and used greedy pruning strategy to only retain variants with p < 5×10^-8^ and r^2^ < 0.01. Although previous studies have used more relaxed r^2^ thresholds for LD pruning (*11*), we found that our high statistical power required a more stringent filtering to avoid including many variants with low residual LD. We performed the primary *cis*-MR analysis using two pleiotropy-robust methods that we found to perform well in the *cis*-MR context in our previous benchmark study (*89*): multiplicative random-effects IVW-MR implemented in the MendelianRandomization R package (*88*) and MRLocus (*90*). For the IVW-MR method, we also specified ‘weights = delta’. We also repeated the same analysis using MR-Egger (*91*), To further assess the robustness of our MR results, we also tested two other *cis*-MR methods: MR-link-2 (*92*) and MR-PCA (*93*). The advantage of these methods is that instead of requiring instrument selection via LD pruning, they explicitly model the LD between all associated variants in the *cis* region. For MR-link-2 and MR-PCA, all association summary statistics were harmonized to the UK10K genotype reference (*94*) and both methods were using the default parameters provided by the ‘mr_link_2_standalone.py function’: MAF > 0.005, regional definition +/- 250kb, and instrument selection threshold p < 5×10^-8^ and variance explained of the LD matrix = 99%.

### Structural modelling

Models of the three subunits of the BCKD complex in Figure 3b were generated using AlphaFold 3 via the AlphaFold Server (*95*). Structures of the regulatory proteins BCKDK and PPM1K were retrieved from the AlphaFold Protein Structure Database (*96, 97*). For clarity in visualization, disordered or unstructured regions at the N- and C-termini were manually removed. Molecular graphics were performed with UCSF ChimeraX (*98*).

## Data and code availability

Complete genetic ancestry group-specific and meta-analysis association summary statistics from this study can be downloaded from the GWAS Catalog (*62*) (accessions GCST90449363 - GCST90451603, Table S12). GWAS lead variants are available from Zenodo (https://dx.doi.org/10.5281/zenodo.13937265). The meta_EUR meta-analysis results can also be viewed in our PheWeb browser (https://nmrmeta.gi.ut.ee/). Meta-analysis code is available from https://github.com/ralf-tambets/EstBB-UKBB-metaanalysis/. The individual-level UK Biobank data are available for approved researchers through the UK Biobank data-access protocol (https://www.ukbiobank.ac.uk/enable-your-research/apply-for-access). The individual-level data from Estonia Biobank can be accessed through a research application to the Institute of Genomics of the University of Tartu (https://genomics.ut.ee/en/content/estonian-biobank).

## Author contributions

R.T. performed GWAS analysis on the EstBB and UKBB data. I.R. developed the initial GWAS workflow. N.T., A.K., K.F. developed quality control criteria for the EstBB metabolite data. R.T. and K.A. designed the *cis*-MR and genome-wide MR analyses and interpreted the results. AvdG performed the MR-link-2 and MR-PCA analyses. M.J. performed genetic colocalisation between molecular QTLs, metabolic traits and diseases. E.A. performed structural modelling of the BCKDH complex. D.Y. obtained SpliceAI and Pangolin predictions for all lead variants. T.E. established the NMR dataset within the EstBB. Z.K., T.E., K.A. and P.P. supervised the research. K.A., P.P., R.T, U.V., E.A. and J.K. wrote the manuscript with feedback from all authors.

## Data Availability

Complete genetic ancestry group-specific and meta-analysis association summary statistics from this study can be downloaded from the GWAS Catalog (62) (accessions GCST90449363 - GCST90451603, Table S12). GWAS lead variants are available from Zenodo (https://dx.doi.org/10.5281/zenodo.13937265). The meta_EUR meta-analysis results can also be viewed in our PheWeb browser (https://nmrmeta.gi.ut.ee/). Meta-analysis code is available from https://github.com/ralf-tambets/EstBB-UKBB-metaanalysis/. The individual-level UK Biobank data are available for approved researchers through the UK Biobank data-access protocol (https://www.ukbiobank.ac.uk/enable-your-research/apply-for-access). The individual-level data from Estonia Biobank can be accessed through a research application to the Institute of Genomics of the University of Tartu (https://genomics.ut.ee/en/content/estonian-biobank).

https://dx.doi.org/10.5281/zenodo.13937265

https://github.com/ralf-tambets/EstBB-UKBB-metaanalysis/blob/main/data/sumstats_paths.tsv

## Acknowledgements

We want to acknowledge the participants of the UK Biobank and Estonian Biobank for their contributions. The Estonian Genome Center analyses were partially carried out in the High Performance Computing Center, University of Tartu. We thank Eric B Fauman for feedback on an earlier version of the manuscript. This research has been conducted using the UK Biobank Resource under application numbers 91233 and 30418. Nightingale Health Plc is acknowledged for early access to the UK Biobank NMR metabolite data. UCSF ChimeraX was developed by the Resource for Biocomputing, Visualization, and Informatics at the University of California, San Francisco, with support from National Institutes of Health R01-GM129325 and the Office of Cyber Infrastructure and Computational Biology, National Institute of Allergy and Infectious Diseases. The Estonian Biobank Research Team consists of Mari Nelis, Georgi Hudjasov, Reedik Mägi, Andres Metspalu, and Lili Milani.

## Supplementary Figures

**Figure S1.**
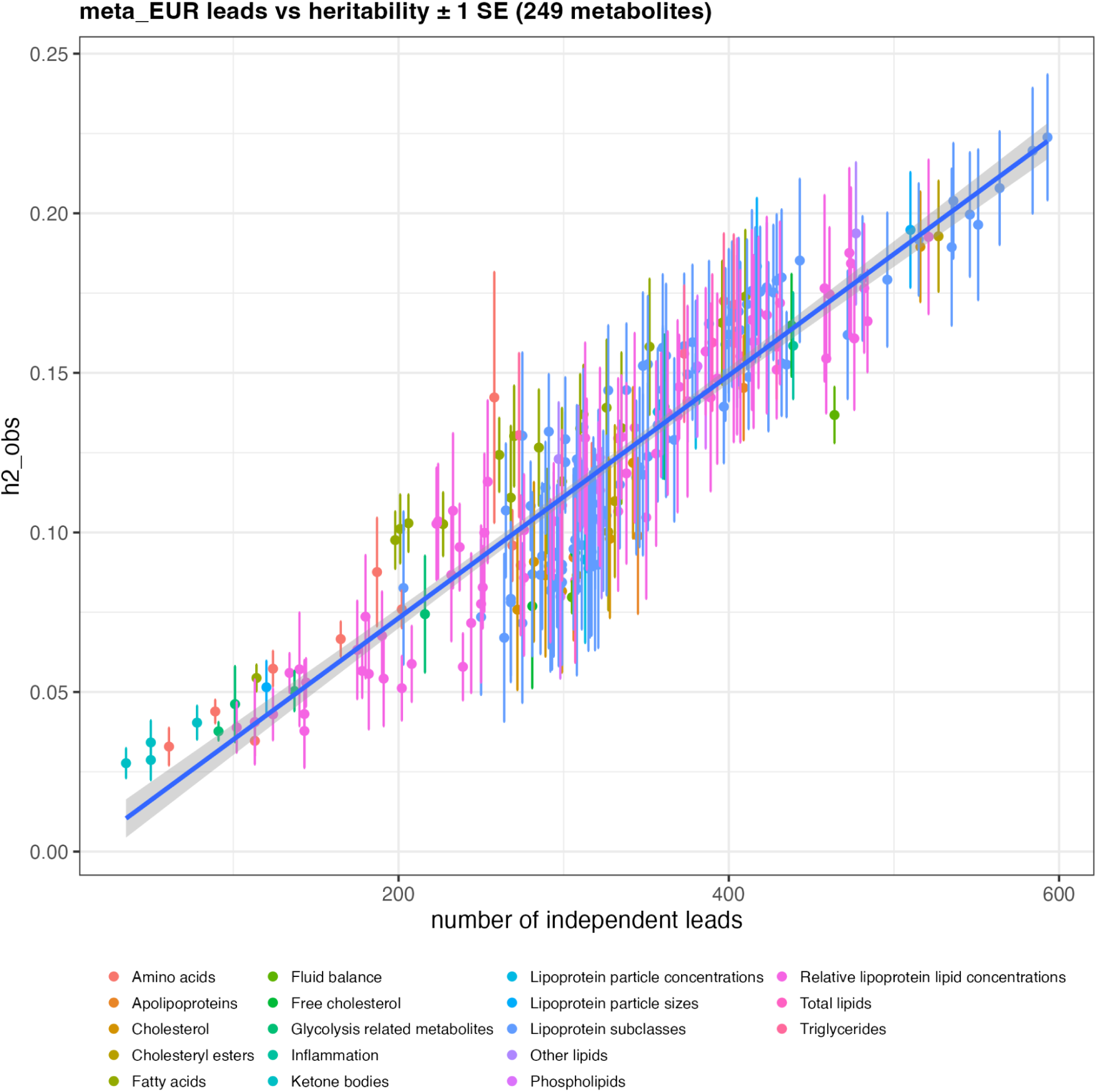
Relationship between heritability (h2_obs) and the number of genome-wide significant hits detected for each of the 249 metabolic traits in the meta_EUR meta-analysis.

**Figure S2.**
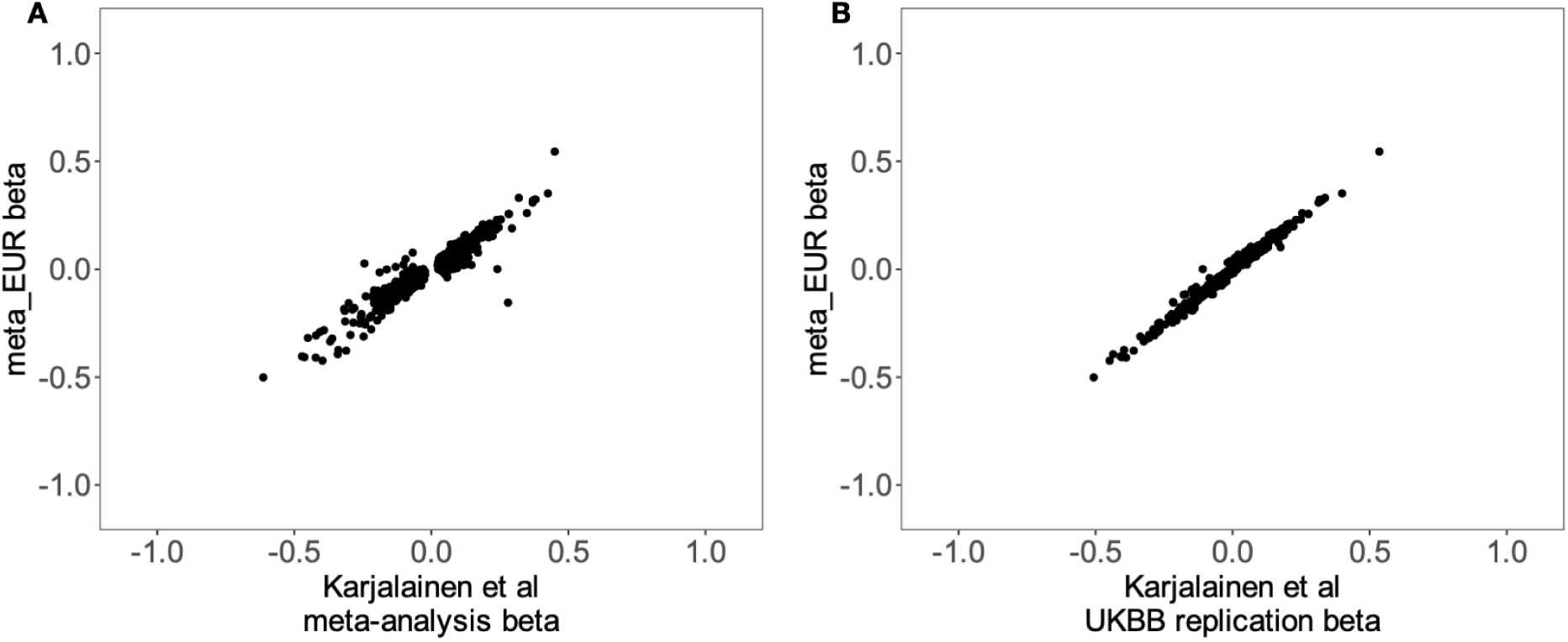
Comparison of shared lead variant betas in our European-ancestry meta-analysis (meta_EUR) and results presented by Karjalainen *et al*. (**A**) Scatter plot of GWAS lead variant effect sizes from Karjalainen *et al* main analysis (n = 137k, 33 cohorts) and our meta_EUR. (**B**) GWAS lead variant effect sizes from Karjalainen *et al* UK biobank replication (n = 100k) and our meta_EUR meta-analysis. Even though Karjalainen *et al* included 3,701 samples from the Estonian Biobank, these were older samples profiled in 2011- 2012 that were excluded from our meta-analysis due to significant batch effects. Thus, there is no sample overlap between our meta-analysis and the primary analysis conducted by Karjalainen *et al* (panel **A**). The ∼100,000 UK Biobank samples used for replication by Karjalainen *et al* were also part of our meta-analysis, explaining the extremely high concordance in GWAS effect sizes.

**Figure S3.**
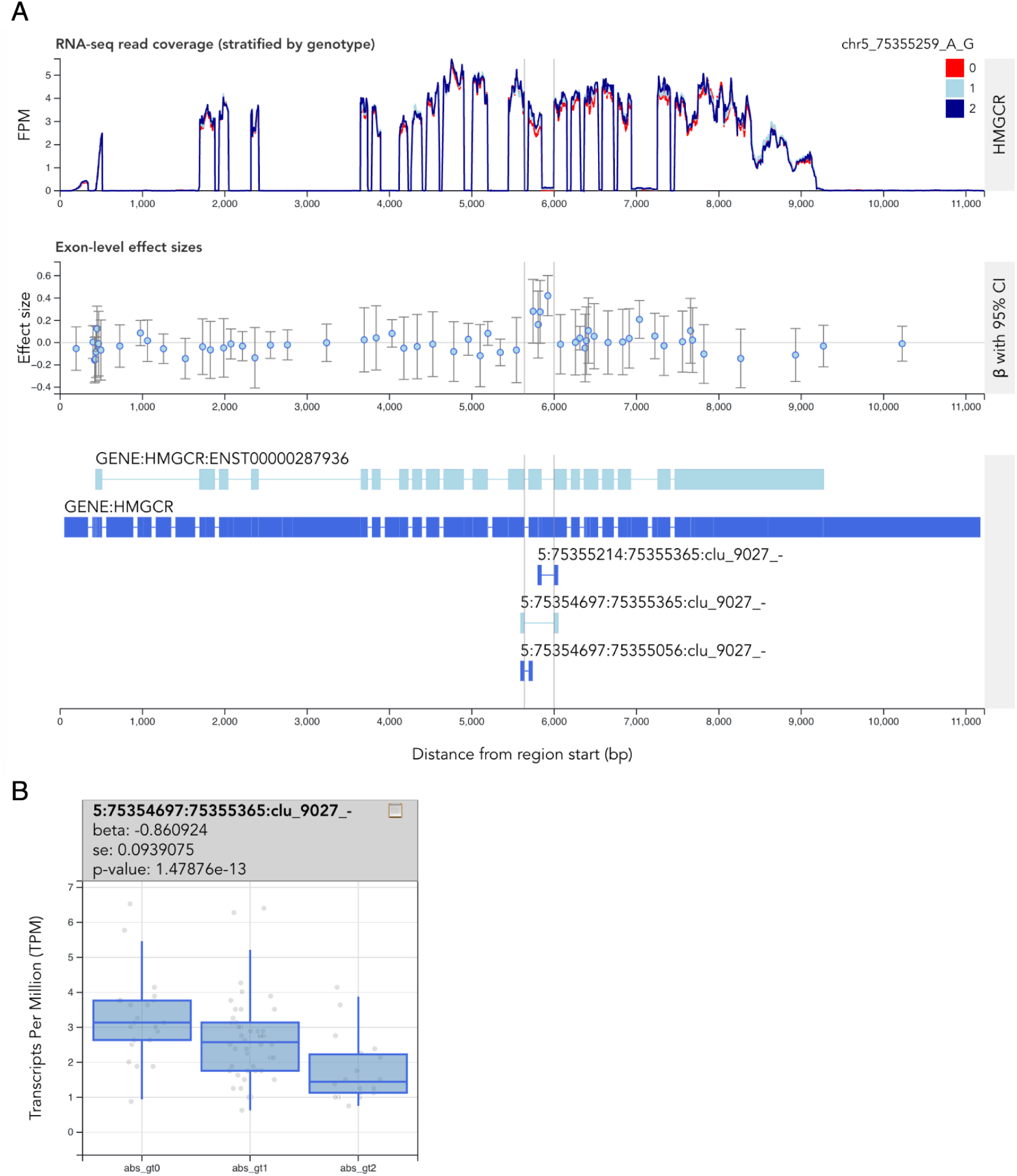
*HMGCR* sQTL signal in the Alasoo_2018 dataset. (**A**) RNA-seq read coverage across the HMGCR gene stratified by the genotype of the lead sQTL variant (5-75355259-A-G). (**B**) Usage of the exon13-skipping splice junction stratified by the genotype of the lead sQTL variant. Interactive visualisation available from <u>here</u>.

**Figure S4.**
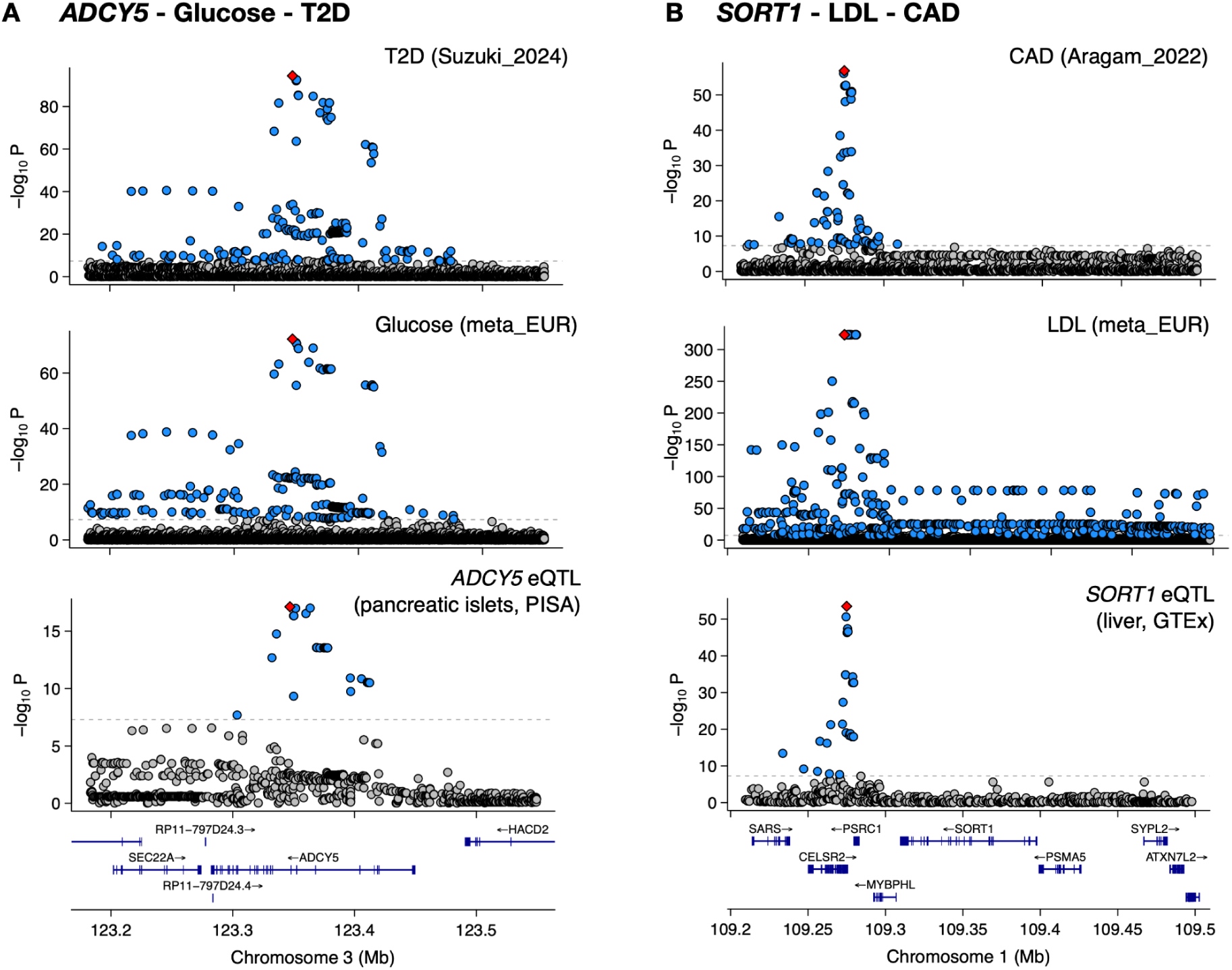
Examples of colocalising eQTL - metabolic trait - disease triplets. (**A**) Colocalisation between *ADCY5* eQTL in pancreatic islets, plasma glucose and T2D GWAS. (**B**) Colocalisation between SORT1 eQTL in the liver, plasma LDL cholesterol and CAD.

**Figure S5.**
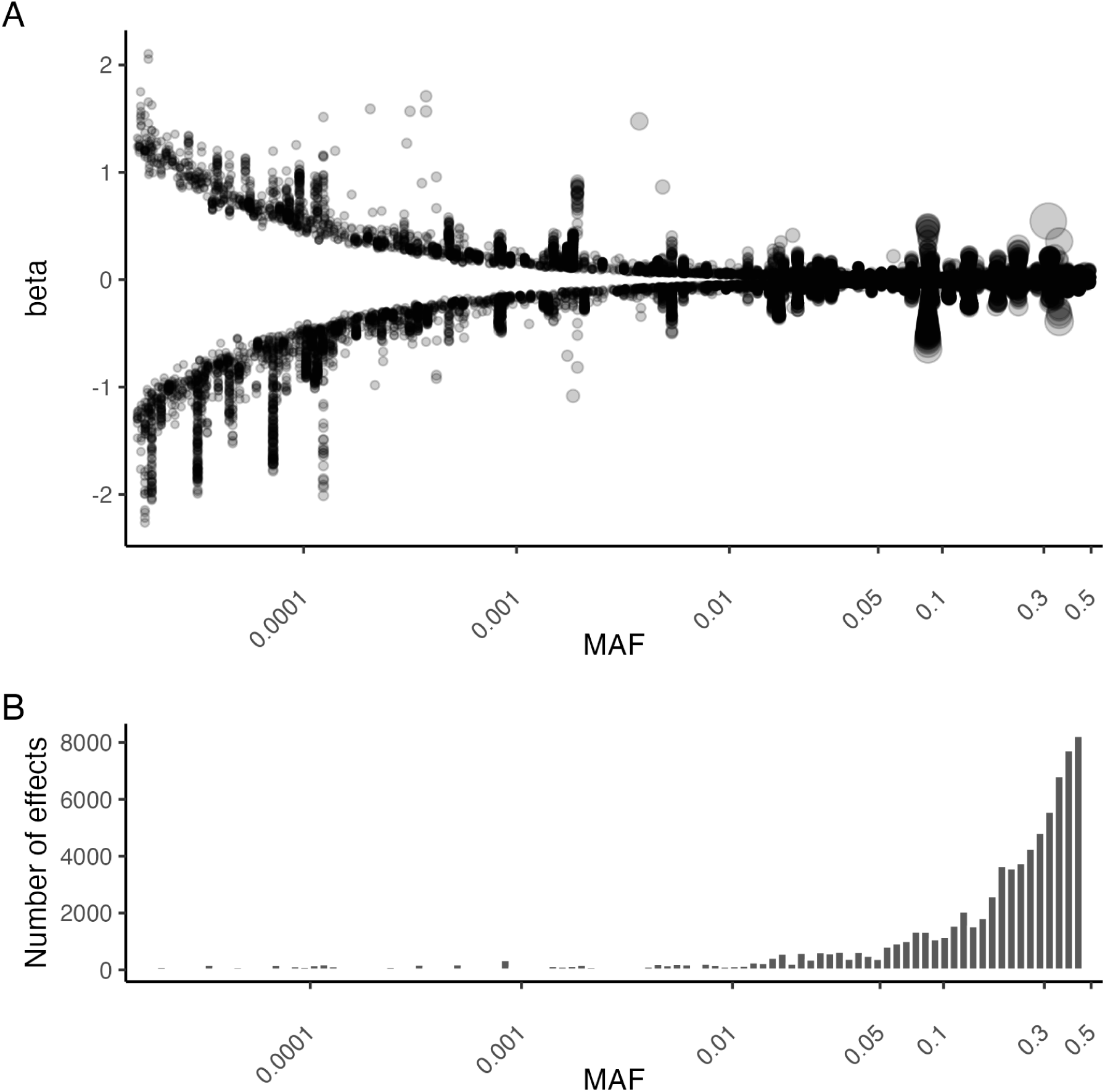
Detected metabolite trait associations with low-frequency variants. (**A**) Relationship between the lead variant minor allele frequency (MAF) and effect size (beta). Each dot signifies the lead variant (+/- 1Mb window) from each locus-trait pair (meta_EUR). The size of each dot has been scaled by -log_10_ p-value. (**B**) Number of detected significant associations in relation to the lead variant MAF in meta_EUR analysis.

**Figure S6.**
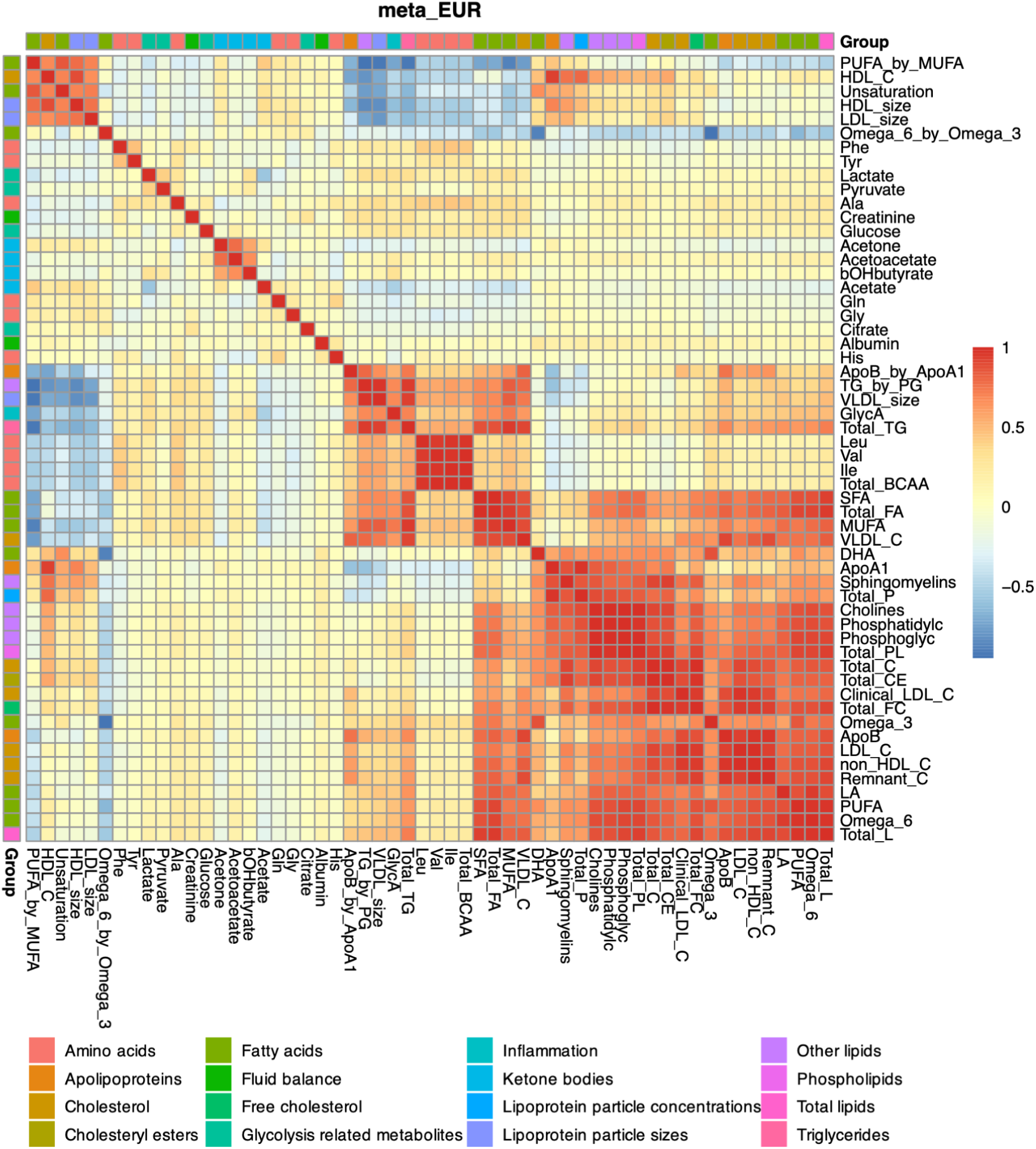
Heatmap of pairwise genetic correlations between metabolic traits in the meta_EUR dataset. The heatmap shows a representative subset of 56 metabolic traits from the main metabolic classes. The complete genetic correlation matrix for all 249 metabolic traits is presented in Table S7.

**Figure S7.**
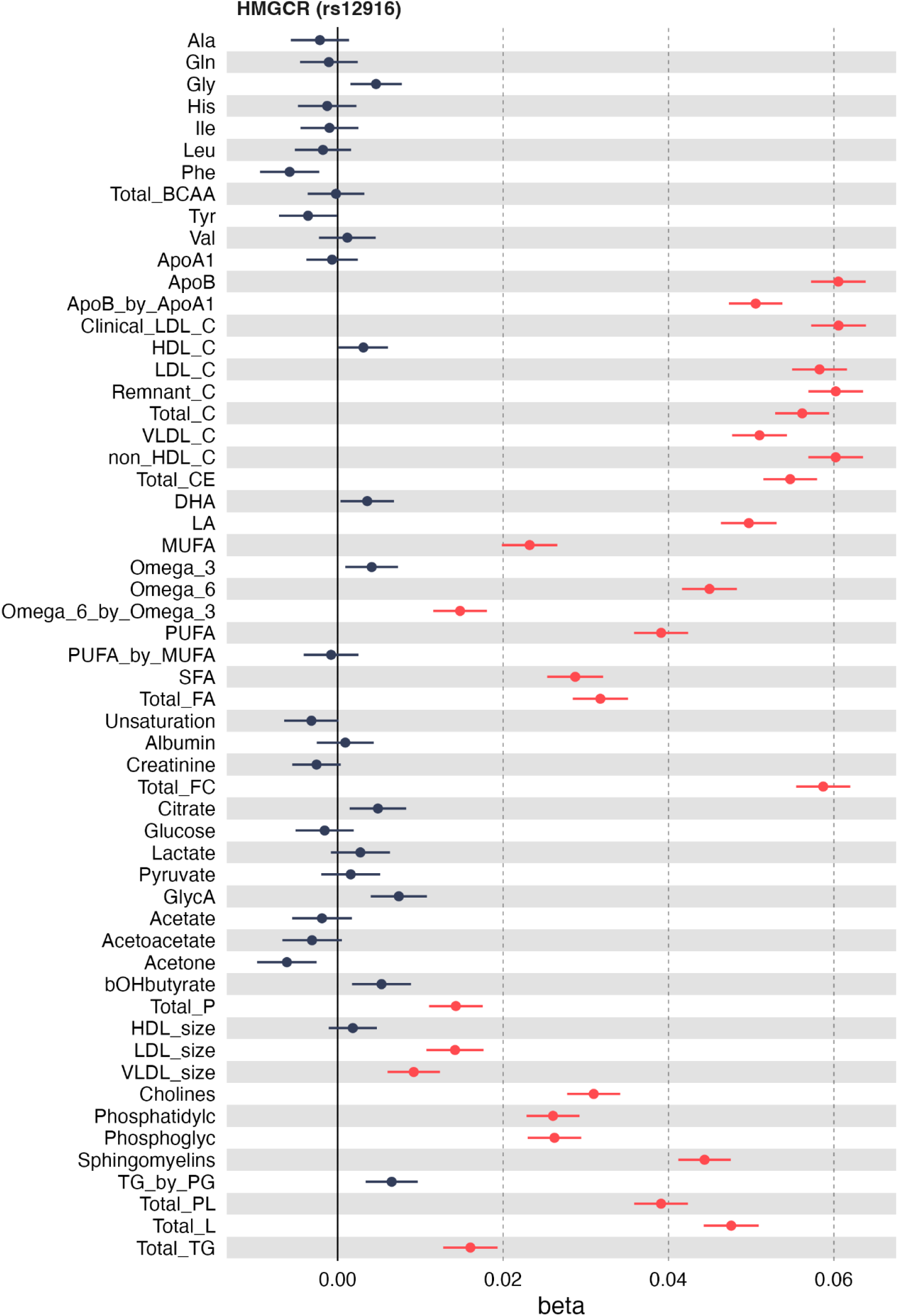
Pleiotropic effects of the *HMGCR* locus lead variant rs12916 on many lipid-related metabolites. The forest plot shows a representative subset of 56 metabolic traits from the main metabolic classes.

**Figure S8.**
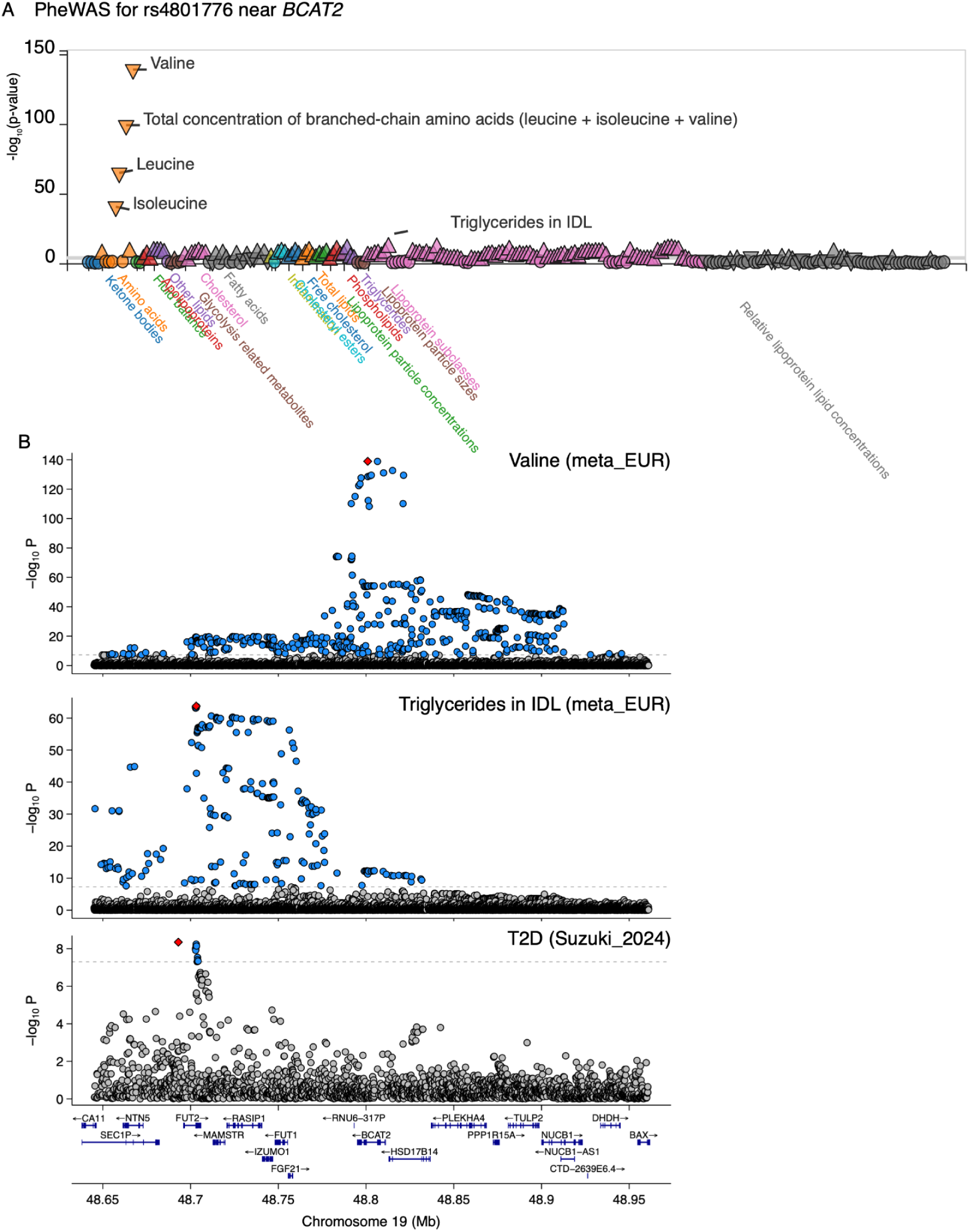
Association signals for BCAAs, lipoprotein traits and T2D near *BCAT2*. (**A**) PheWAS plot for the valine lead variant (rs4801776) in the intron of *BCAT2*. In addition to very strong associations with all three branched-chain amino acids, we also see genome-wide significant associations with various lipid traits. (**B**) Regional association plots for Valine, Triglycerides in IDL and T2D in the *BCAT2* region. The association between rs4801776 and lipid traits seems to be driven by an independent lipid signal near the *FUT2* gene that has low LD (r^2^ = 0.08) with the valine lead variant.

**Figure S9.**
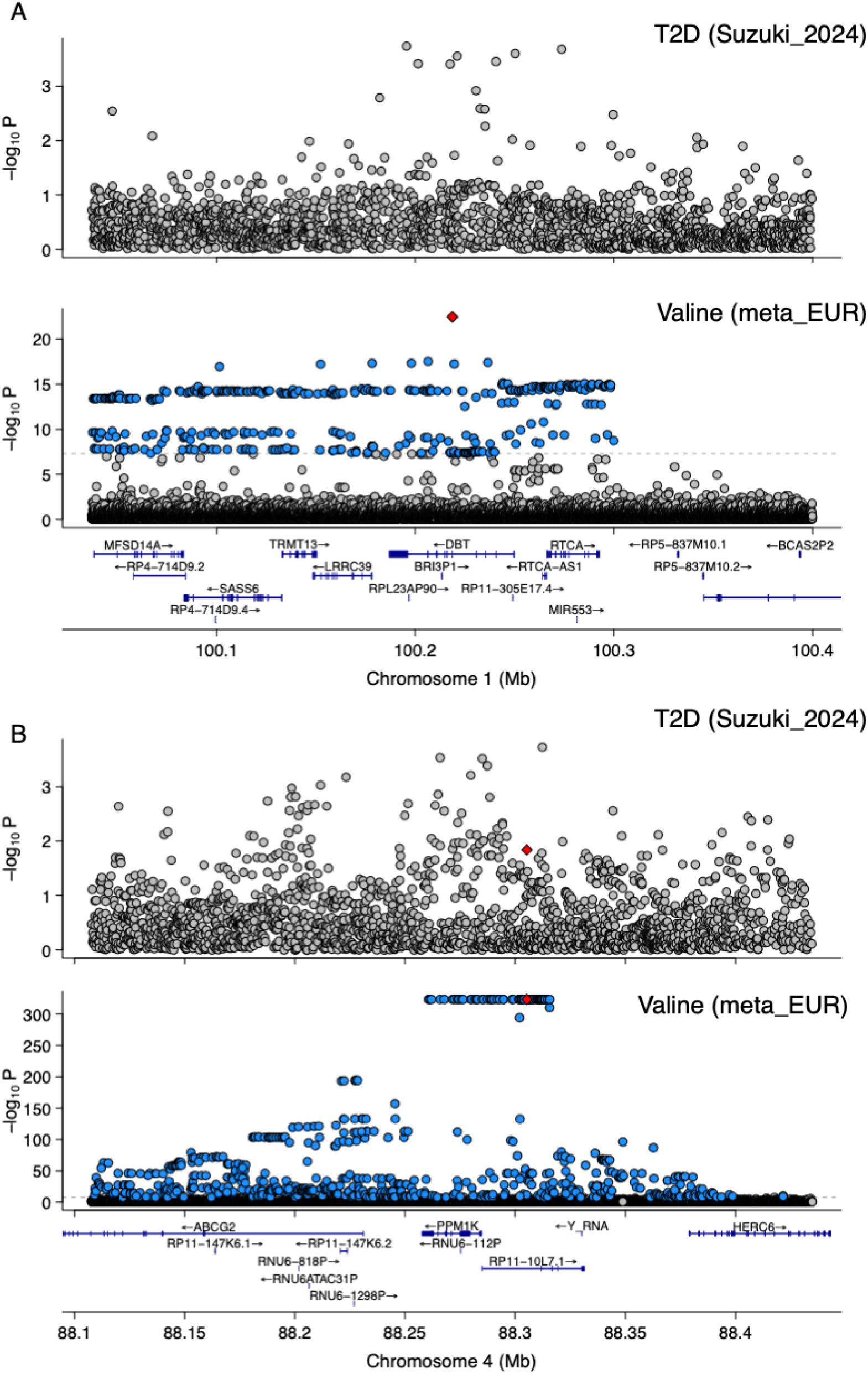
Regional association plots for T2D and valine in the *cis* regions of (**A**) *DBT* and (**B**) *PPM1K*.

